# Comparative evaluation of EPI and SWI for the assessment of PRL and CVS in Multiple Sclerosis

**DOI:** 10.64898/2026.02.05.26345463

**Authors:** Anna Stölting, Eline van Doninck, Serena Borrelli, Colin Vanden Bulcke, Maria Sofia Martire, Francois Guisset, Maxence Wynen, Gaëtan Duchêne, Lucia Moiola, Veronica Popescu, Barbara Willekens, Massimo Filippi, Martina Absinta, Pietro Maggi

## Abstract

**Introduction:** The 2024 McDonald criteria incorporate the central vein sign (CVS) and paramagnetic rim lesions (PRL) as supportive imaging biomarkers for MS diagnosis. While susceptibility-weighted-imaging (SWI) and T2*-weighted echo-planar-imaging (EPI) are generally used to assess CVS/PRL, their relative performance remains unclear. This study compared high-resolution isotropic-T2*-EPI with non-isotropic SWI for CVS/PRL detection.

**Materials and Methods:** In this multi-centre study, 21 patients with MS underwent harmonized 3T-MRI including EPI and SWI. CVS and PRL were evaluated according to NAIMS criteria. Whole-brain and controlled lesion analyses on 120 pre-selected lesions were performed independently for each contrast, with EPI serving as reference standard.

**Results:** In whole-brain analyses, SWI showed good sensitivity for CVS eligibility and positivity (AC1=0.68-0.78) but significant directional disagreement with EPI (p<0.0001). Discrepancies were primarily attributed to limited lesion-parenchyma contrast and venous visibility on SWI, which improved using low-flip-angle SWI. Controlled lesion analyses supported these observations. For PRL, SWI demonstrated high sensitivity (88%) and precision (97%) compared to EPI, though systematic bias persisted (p<0.001). Controlled lesion analyses showed more balanced, albeit moderate performance.

**Conclusion:** SWI diverged systematically from EPI for CVS and PRL detection. When available, EPI should be preferred, while optimised low-flip-angle SWI may serve as an alternative to conventional SWI.

## Introduction

The 2024 update to the McDonald MS diagnostic criteria^1^ incorporated the central vein sign (CVS) and presence of paramagnetic rim lesions (PRL) as imaging markers supporting the diagnosis of multiple sclerosis (MS). This reflects the growing evidence for their diagnostic and prognostic relevance in MS.^2-4^ Although several magnetic resonance imaging (MRI) sequences have been proposed for detecting these biomarkers in research settings,^5,6^ it remains uncertain which sequence provides the most reliable and consistent visualization in clinical practice. Because both CVS and PRL have recently gained importance in routine diagnostic workflows, identifying the most reliable and practical sequence is of immediate clinical relevance.

The central vein sign reflects the perivenular origin of MS inflammatory demyelinating lesions,^7^ and demonstrates high specificity in differentiating MS from a range of radiological mimics.^8-10^ A substantial proportion of MS lesions contain a central vein sign,^11,12^ and simplified diagnostics paradigms such as “select 3”^13,14^ and “select 6”^9,13^ CVS-positive lesions have shown high sensitivity and specificity for distinguishing MS from non-MS disorders.^15^ Several imaging approaches have been used to visualise the central vein, including susceptibility-weighted imaging (SWI),^16-18^ 3D T2*-weighted echo-planar imaging (EPI),^5^ and combined reconstructions such as FLAIR* or FLAIRswi, which combine susceptibility weighted and FLAIR information.^12,19^ Only a limited number of studies have directly compared the performance of these sequences, though available evidence suggests slightly higher accuracy for high resolution 3D T2*-EPI, as well as for certain combination images.^20-25^

PRL represent another highly specific susceptibility-based MS imaging biomarker. Here, the characteristic paramagnetic rim visible on susceptibility-based MRI,^3,26^ is mainly due to the accumulation of iron-laden microglia and macrophages along the lesion edge.^26,27^ PRL have been associated with poorer clinical outcomes, including faster disability progression and greater disability burden.^4,26,28,29^ While PRL detection is most reliable at high-field MRI, recent studies have demonstrated good visualisation at lower field strengths as well.^30,31^ Importantly, PRL identification requires susceptibility-based imaging techniques,^32^ yet it remains unclear which specific sequence provides the optimal compromise between sensitivity, specificity, and practical clinical use.

Although SWI is widely available in clinical MRI protocols, it has well-recognised limitations for venous assessment, including anisotropic resolution, orientation-dependent susceptibility effects and acquisition times.^16-18^ By contrast, high-resolution 3D T2*-EPI provides isotropic coverage and strong venous conspicuity,^1,5,20^ but is less frequently implemented in routine clinical workflows despite its demonstrated advantages. Given the increasing diagnostic importance of CVS and PRL, determining which susceptibility-based sequence most reliably captures venous structures and chronic active lesions is essential for standardized clinical application.

In this study, we compare high-resolution 3D isotropic T2*-EPI with high-resolution 3D non-isotropic SWI to evaluate their respective performance in detecting CVS and PRL.

## Materials and Methods

### Participants

Clinical and imaging data were collected under institutional review board-approved protocols in two university hospitals (Cliniques universitaires Saint-Luc in Brussels, Belgium and Multiple Sclerosis Center, IRCCS San Raffaele Hospital in Milan, Italy) from March 2022 to July 2025. These protocols allowed pooling of anonymized data. Clinical information matched to MRI data included the Expanded Disability Status Scale,^33^ as assessed by experienced MS clinicians.

All participants provided written informed consent prior to the study enrolment. Study inclusion criteria included: 1) age ≥ 18years, 2) diagnosis of MS according to 2024 McDonald MS Criteria^1^, availability of matched 3T high-resolution 3) echo-planar imaging,^5^ and 4) susceptibility weighted imaging (Figure 1).^34^

**Figure 1.**
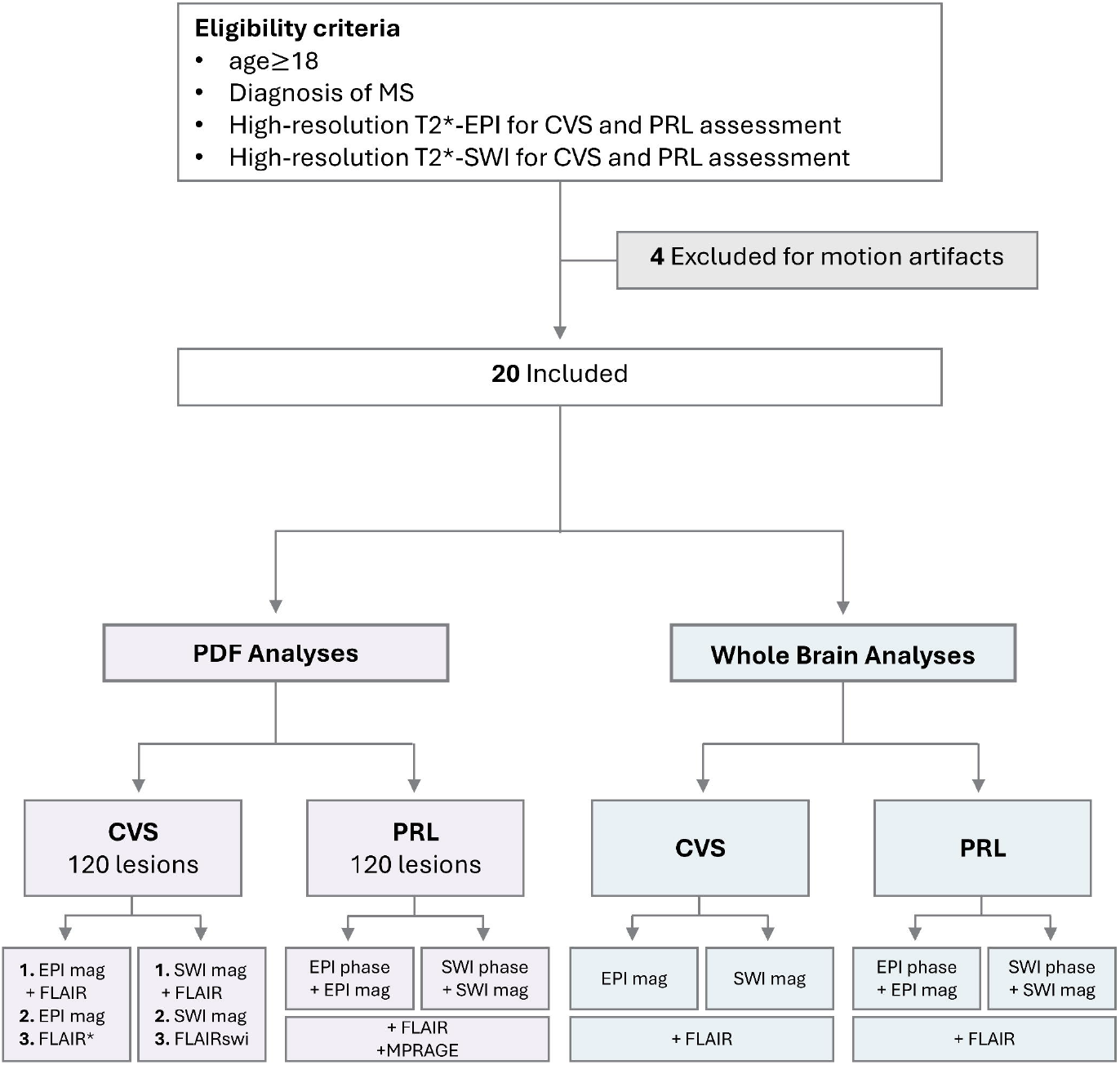
Experimental Workflow Participants were included after fulfilling all inclusion criteria. For controlled lesion analyses of the central vein sign (CVS), 120 pre-selected lesions were evaluated across three experiments, each comprising 40 lesions and presented in standardized PDF files. In Experiment 1, lesions were shown using either EPI magnitude images or SWI magnitude images, with FLAIR images co-registered to the respective susceptibility modality. Experiment 2 presented lesions using susceptibility images only (EPI magnitude or SWI magnitude), without FLAIR. In Experiment 3, lesions were presented using combined FLAIR* or FLAIRswi images. Image presentations were separated by at least 4 weeks to ensure blinding to previous ratings. For controlled lesion analyses of paramagnetic rim lesions (PRL), 120 lesions were pre-selected and presented in standardized PDF files containing FLAIR and MPRAGE images co-registered to either EPI or SWI, together with the corresponding magnitude and phase images. All lesions presented in the controlled analyses included scale rulers for reference. For the whole-brain analyses, raters evaluated CVS using EPI magnitude or SWI magnitude images with FLAIR co-registered to the respective susceptibility modality, and PRL using combined phase and magnitude images (EPI or SWI) with FLAIR co-registered accordingly. EPI- and SWI-based assessments were performed in temporally separated sessions (≥4 weeks apart) to maintain blinding.

Cohort demographics are summarized in Table 1.

### MRI imaging acquisition

MR imaging was performed on a 3T SIGNA™ scanner (GE Health Care) in Brussels, Belgium or Philips Ingenia 3T CX MR in Milan, Italy. The imaging protocol in both centres included a 3D Fluid-attenuated inversion recovery (FLAIR), magnetization prepared rapid gradient echo images (MPRAGE)^35^ and high-resolution 3D T2* EPI and matched SWI, post-gadolinium contrast T1-weighted imaging was used to verify absence of gadolinium-enhancing lesions.

In Brussels, a high-resolution 3D T2*-weighted EPI sequence was acquired in the sagittal plane (TR = 80.2 ms, TE = 35 ms, flip angle = 18°, #slices = 355, voxel size = 0.67 × 0.67 × 0.67 mm^3^, acquisition time = 3:59min). In addition, a 3D axial susceptibility sequence (GE SWAN) was acquired, providing T2*-weighted magnitude images and unwrapped filtered phase contrasts (TR = 50 ms, TE = 20.0 ms, flip angle = 15°, #slices = 96, acquisition voxel size = 0.5 × 0.5 × 1.8 mm^3^, acquisition time = 7:08min). Furthermore, an adapted low–flip-angle SWI sequence was acquired in the axial plane (TR = 36.5 ms, TE = 25.6 ms, flip angle = 12°, #slices = 240, acquisition voxel size = 0.75 × 0.75 × 1.0 mm^3^, acquisition time = 5:26min).

In Milan, a high-resolution 3D T2*-weighted EPI sequence was acquired in the axial plane (TR = 50.7 ms, TE = 27.7 ms, flip angle = 10°, #slices = 336, voxel size = 0.5 × 0.5 × 0.55 mm^3^, acquisition time = 3:59min). In addition, a whole-brain 3D axial SWI sequence was acquired, providing T2*-weighted magnitude images and unwrapped filtered phase contrasts (TR = 31 ms, TE = 7.2 ms, flip angle = 17°, #slices = 261, acquisition voxel size = 0.45 × 0.45× 1mm^3^, acquisition time = 6:04min).

Further MRI sequence details can be found in the supplementary material.

### MR image post-processing

Imaging postprocessing was centralized. FLAIR and MPRAGE images were rigidly registered to either EPI magnitude or SWI magnitude using the Advanced Normalisation Tools (ANTs).^36^ This co-registration enabled the assessment of PRL and CVS directly on the native, unprocessed susceptibility images while maintaining anatomical reference.

FLAIR and susceptibility magnitude images were combined to generate FLAIR* or FLAIRswi images.^37^ As SWI data are usually reconstructed with vendor-applied phase filtering during acquisition, we applied an in-house filtering protocol to the EPI phase images to approximate the characteristics of clinically generated SWI images and ensure comparability across centers.^12-14^ EPI phase images were thus filtered with an in-house algorithm using denoising^40^ and dehazing, to enhance image quality and allow fair comparison to the filtered SWI phase.

PRL and CVS were assessed according to the North American Imaging in Multiple Sclerosis (NAIMS) guidelines.^11,41^ Detailed criteria can be found in the supplementary material. Based on these criteria, lesions were classified with a binary rating for PRL (PRL vs. non-PRL) and into one of three categories for CVS: CVS+, CVS-, or CVS non-eligible (CVS-NE).

### Image Analysis

Image analysis comprised two independent components: whole-brain assessments and controlled lesion-based assessments (Figure 1). In whole-brain analyses, four experienced raters independently evaluated CVS (2 raters) and PRL (2 raters) on EPI- and SWI-based images coregistered to FLAIR, with assessments performed in temporally separated experiments (≥4 weeks apart) and final consensus adjudication. Controlled lesion analyses evaluated 120 pre-selected lesions presented in standardized PDFs, with CVS assessed across three contrast scenarios (FLAIR + susceptibility, susceptibility only, and combined FLAIR*/FLAIRswi images) and repeated for EPI and SWI in separate sessions (≥4 weeks apart; 3 raters). PRL were evaluated using phase- and magnitude-based susceptibility imaging with anatomical reference (FLAIR and MPRAGE; 6 raters). Full details on rater assignment, lesion selection, adjudication procedures, and image presentation are provided in the Supplementary Material.

### Statistical Analysis

Lesion-level agreement was evaluated for PRL (PRL vs. non-PRL) and CVS (CVS+, CVS-, CVS-NE) classifications. For both whole-brain and controlled lesion analyses, consensus ratings were generated by majority vote; remaining discrepancies were adjudicated by an independent rater. These consensus masks served as the reference for comparing EPI and SWI ratings. EPI was treated as the operational reference standard because of its higher spatial resolution and 3D isotropic visualisation of the imaging biomarkers under study. Discrepant classifications between EPI and SWI were reviewed, and the underlying causes were documented.

Differences between centres were assessed using the Wilcoxon rank-sum test. Analyses were performed using the mean PRL count or mean number of CVS-eligible lesions across both contrasts as well as separately for each contrast. Lesions were categorized as true positive (TP), true negative (TN), false positive (FP), or false negative (FN) relative to the EPI reference, and sensitivity, specificity, accuracy, and precision were calculated. Agreement across contrasts was quantified using both Cohen’s κ and Gwet’s AC1. But as κ is highly sensitive to prevalence imbalance whereas AC1 provides a more stable estimate under such conditions, we are reporting Cohen’s κ results in the supplementary. McNemar’s test was used to assess paired categorical outcomes. When the number of discordant pairs was <25, the exact McNemar’s test was applied.

## Results

### Whole Brain Analysis

#### CVS Analysis

No centre differences were observed in lesion eligibility, whether assessed using the number of eligible lesions across contrasts or using EPI or SWI alone (all p>0.7).

After consensus, a total of 174 lesions were identified as eligible for the CVS assessment. Of those, 168 (97%) were classified as eligible on EPI and 137 (79%) on SWI. More specifically, 131 lesions (75%) were eligible on both sequences (TP) while 6 (3%) were eligible only on SWI (FP) and 37 (21%) were eligible only on EPI (FN). All FP lesions (100%) exhibited multiple veins on EPI that were not visualized on SWI. Among FN lesions, the majority (n=31, 84%) were deemed non-ratable as the lesion itself was not visible on SWI (Figure 2B), while for the remainder (n=6, 16%) it was impossible to differentiate between the lesion parenchyma and its central vein due to diffuse intralesional hypointensity on SWI magnitude images (Figure 2C). These lesions were subsequently identified as PRL on phase images.^42,43^ Of note, out of the 37 FN lesions, 1 lesion was located in the cerebellum, while 13 (35%) were juxtacortical and 23 (62%) were subcortical, in close proximity to the cortex but not touching it.

**Figure 2.**
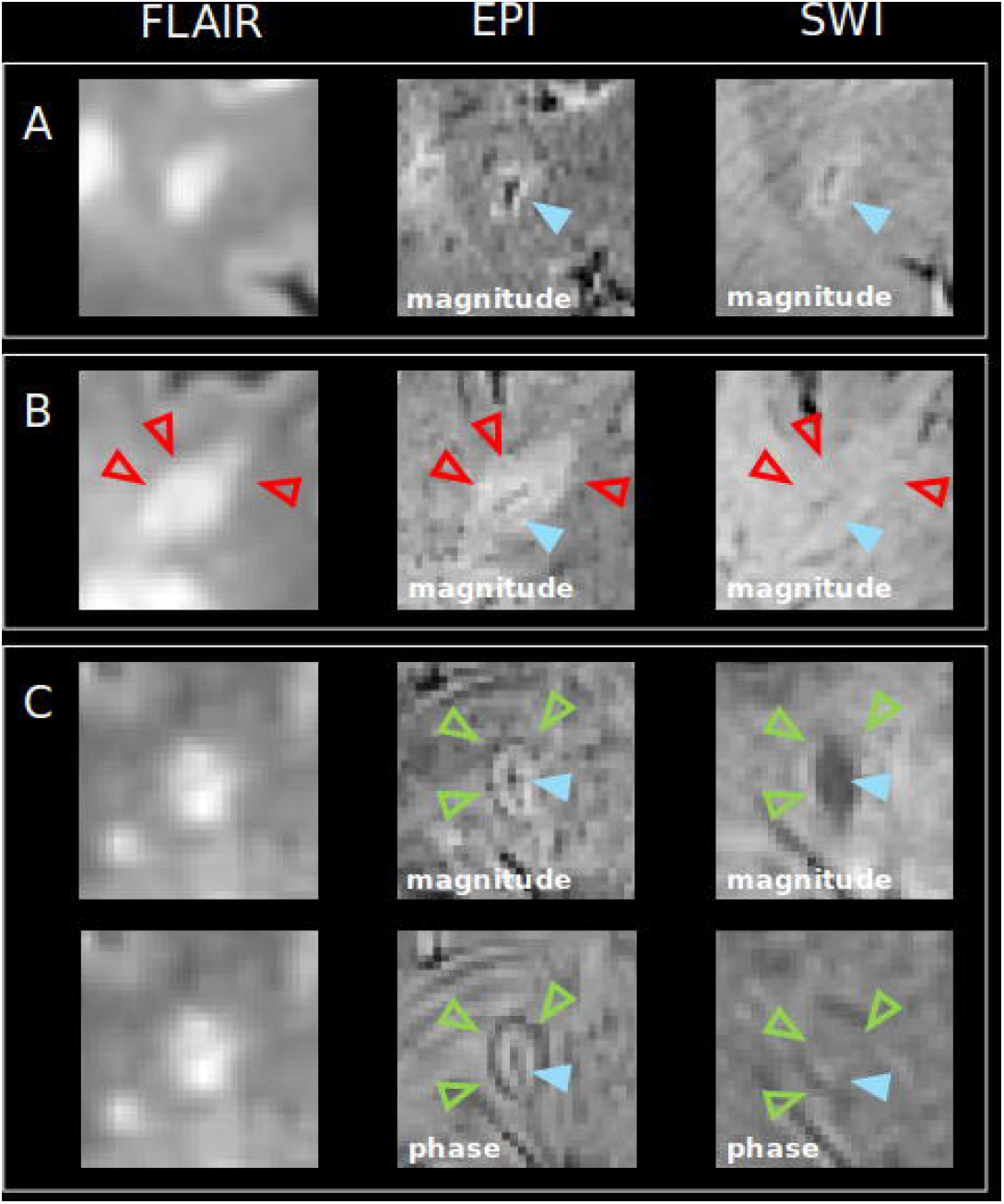
Examples of CVS visualised on EPI and SWI magnitude A: CVS visible on both EPI and SWI magnitude (light blue arrow). B: CVS clearly visible on EPI magnitude (blue arrow) but difficult to distinguish on SWI magnitude due to reduced lesion contrast (red arrows). C: CVS+ PRL (blue and green arrows) visible on EPI magnitude and phase, while neither CVS nor PRL are visible on SWI magnitude or phase. Abbreviations: CVS: central vein sign; EPI: echo-planar imaging; PRL: paramagnetic rim lesion; SWI: susceptibility-weighted imaging.

For eligibility, SWI showed good sensitivity of 78% (95% CI: 0.71-0.84), and accuracy of 75% (95% CI: 0.68-0.82). AC1 indicated substantial agreement (AC1=0.68, 95% CI: 0.58-0.79, p<0.0001), but McNemar’s test demonstrated significant asymmetry in paired ratings (p<0.0001), with EPI classifying more lesions eligible than SWI.

Among the 131 lesions eligible on both contrasts, 104 (79%) were CVS+ on both sequences (TP) (Figure 2A), 23 (18%) were CVS+ on EPI but CVS-on SWI (FN), and 4 (3%) were CVS-on both sequences (TN). Of the 23 FN lesions, 19 (83%) were located subcortically, while 4 (17%) were juxtacortical lesions. For CVS identification, SWI demonstrated good sensitivity of 82% (95% CI: 0.74-0.88), excellent specificity of 100% (95% CI: 0.4-1.0), and good accuracy of 82% (95% CI: 0.75-0.89). Agreement between contrasts was substantial using AC1 (AC1=0.78, 95% CI: 0.68-0.88, p<0.001), but McNemar’s test again indicated significant asymmetry in paired ratings (p<0.0001).

All results are provided in Supplementary Table 2.

##### Adapted low-flip angle SWI

One subject underwent an MRI acquisition using a low-flip angle SWI sequence, with a matched EPI acquisition. This subject was evaluated, and agreement between sequences was reassessed (Figure 3B).

**Figure 3.**
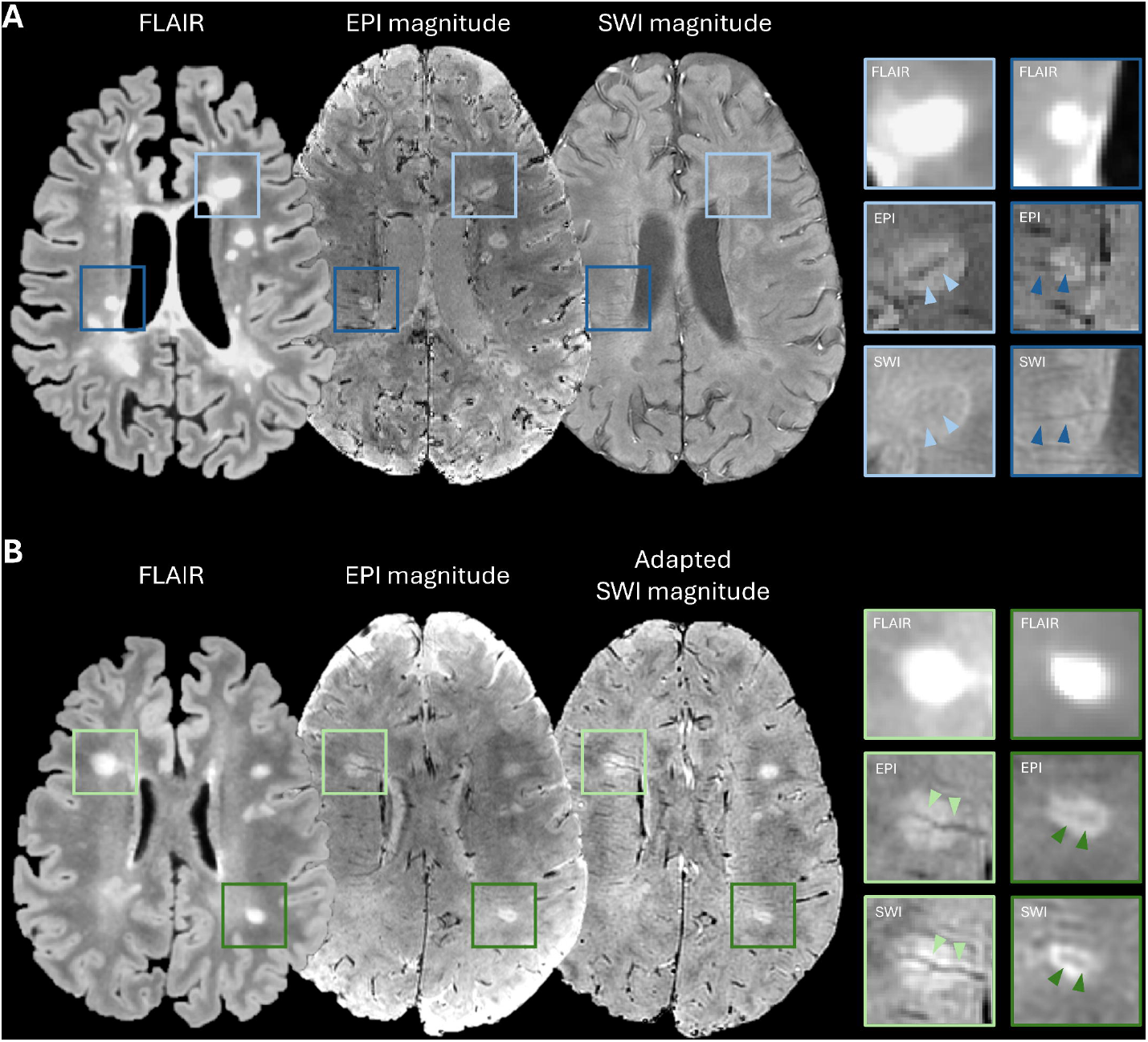
Comparison of standard SWI and adapted SWI A: Representative EPI magnitude and standard SWI magnitude images. The light blue box highlights a lesion in which the central vein is visible on EPI but not on standard SWI. The dark blue box highlights a lesion in which the lesion parenchyma is not visible on standard SWI, although a central vein can still be identified. B: Representative EPI magnitude and adapted SWI magnitude images. The light green and dark green boxes highlight lesions in which both the lesion parenchyma and the central vein are clearly visible on both contrasts.

Fifteen lesions were identified on EPI, of which fourteen were also identifiable on SWI. One FN lesion was only rateable on EPI, as it was virtually invisible on SWI (sensitivity 93%). Specificity was not estimable because no EPI-negative lesions were included. Agreement for lesion eligibility was high (Gwet’s AC1= 0.93, p<0.0001), and McNemar’s test showed no evidence of directional asymmetry (p=1.0). Among the fourteen lesions visible on both contrasts, no discrepancies in CVS classification were observed: all lesions were rated as CVS-positive on both EPI and SWI.

#### PRL Analysis

The median number of PRL did not differ between centres (p = 0.38). Similar results were obtained when analysing EPI-derived PRL counts alone (p = 0.32) and SWI-derived PRL counts alone (p = 0.34).

Out of the 311 identified PRL, 265 lesions (85%) were rated PRL on both contrasts (TP) (Figure 4A), 37 lesions (12%) were PRL on EPI but non-PRL on SWI (FN), and 9 lesions (3%) were PRL on SWI but not on EPI (FP). Among the 37 FN lesions, 25 (68%) were entirely invisible on SWI (Figure 4B), 3 (8%) did not meet the two-thirds closed-rim criterion, and 9 (24%) showed insufficient differentiation between the hypointense rim and lesion parenchyma, thereby failing to meet the closed rim criterion.^41^ Of the 9 FP lesions, 1 exhibited rim-mimicking veins, while 8 (89%) were PRL on SWI but not on EPI (the latter category corresponding to PRL located in regions affected by susceptibility artifacts on the EPI, see below).

**Figure 4.**
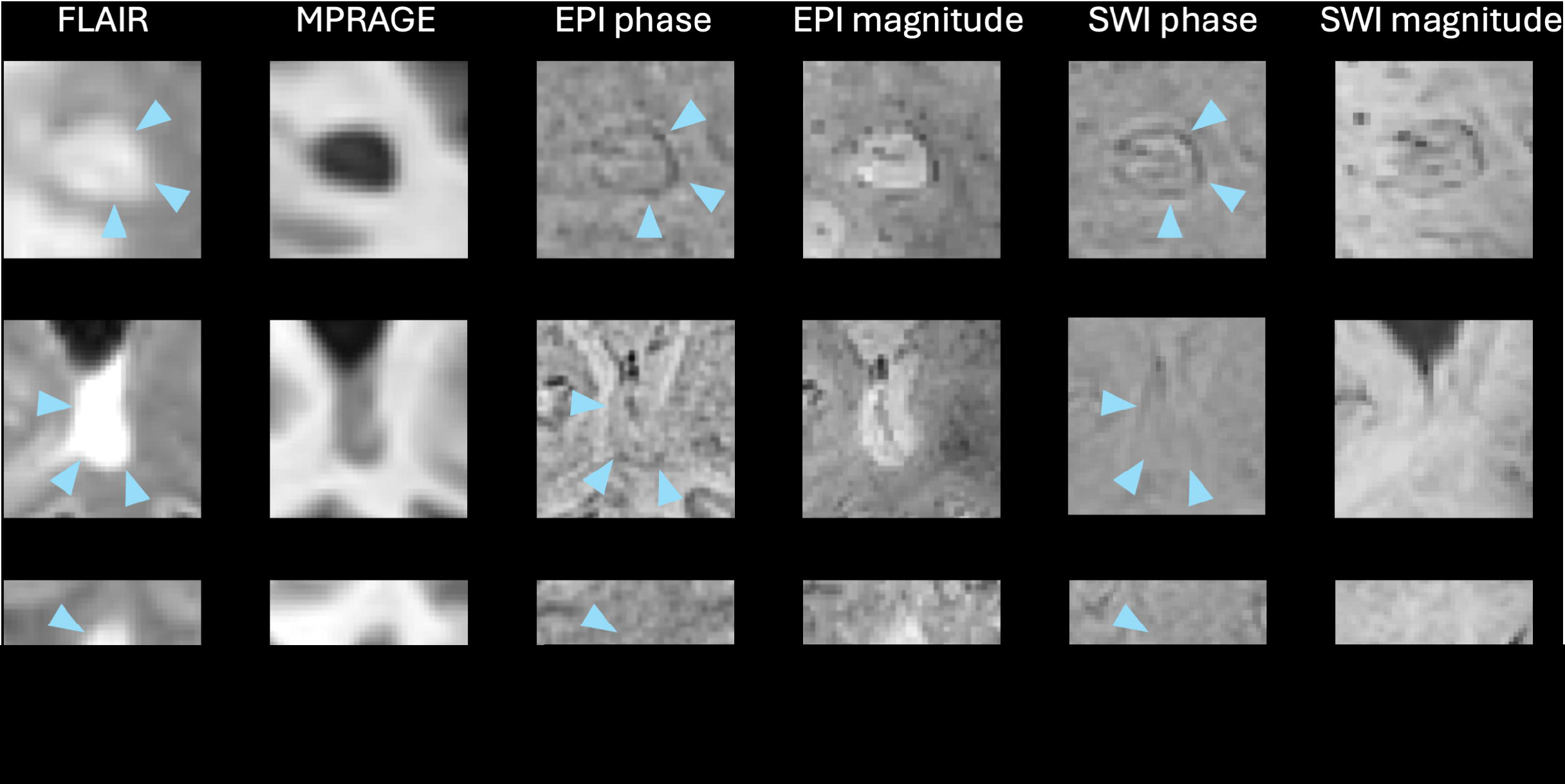
Examples of PRL visualised on EPI phase and SWI phase images A: PRL visible on both EPI and SWI phase with prominent central vein sign on both EPI and SWI magnitude. B: PRL visible only on EPI phase but not SWI phase. Lesion located at the posterior horn of the lateral ventricle. C: non-PRL on both EPI and SWI phase.

Anatomically, 34 of the 37 FN lesions (92%) were located either in the deep white matter (WM; n=19, 56%) or periventricular regions (n=15, 44%), with the remaining 3 lesions (8%) located juxtacortically. Within the deep WM, 7 lesions were situated near the cortex and 4 along the optic pathway. Among the periventricular FN lesions, 8 were located around the body of the lateral ventricle, 5 near the occipital horn, and 2 at the temporal horn. The 9 FP lesions were distributed across deep WM (n=3), periventricular regions (n=3), juxtacortical areas (n=1), and supratentorial regions (n=2). Among the periventricular FP lesions, 2 were located around the body of the lateral ventricle and 1 at the anterior horn. Both supratentorial FP lesions were in the cerebellum. Notably, all FP lesions corresponded to regions on EPI that were difficult to assess due to susceptibility-induced geometric distortions and signal dropout.^44,45^

For PRL detection, SWI demonstrated good sensitivity of 88% (95% CI: 0.84-0.91) and excellent precision of 97% (95% CI: 0.94-0.98). Specificity could not be meaningfully estimated due to the absence of TN lesions; the resulting value of 0% reflects the structure of the sample rather than true diagnostic performance. Overall accuracy was good at 85% (95% CI: 0.81-0.89). Agreement analysis indicated good agreement (AC1=0.82, 95% CI: 0.78-0.88, p<0.001), although McNemar’s test was significant (p<0.001), confirming systematic bias, with EPI classifying substantially more lesions as PRL than SWI.

All results are provided in Supplementary Table 1.

### Controlled Lesion Analysis

#### CVS Analysis

Controlled lesion analysis was divided in three sub-experiments where raters were presented with different image combinations.

##### FLAIR + Susceptibility contrast (EPI magnitude or SWI magnitude)

In Experiment 1 (Figure 1), 18 of 40 lesions (45%) were eligible on both contrasts (TP), 3 lesions (7.5%) were non-eligible on EPI but eligible on SWI (FP), 14 lesions (35%) were non-eligible on both (TN), and 5 lesions (13%) were eligible on EPI but not on SWI (FN). All FN lesions were invisible on SWI, while FP lesions appeared multi-veined on EPI but not on SWI.

SWI demonstrated substantial sensitivity of 78% (95% CI: 0.56-0.92) and specificity of 82% (95% CI: 0.56-0.96). Accuracy was 80% (95% CI: 0.64-0.91). Agreement for lesion eligibility was moderate according to AC1 (AC1=0.60, 95% CI: 0.35-0.86, p<0.001). McNemar’s test was nonsignificant (p=0.73), indicating no systematic directional bias.

Among the 18 eligible lesions, 11 (61%) were CVS+ on both contrasts (TP), 5 (28%) were CVS-on both (TN), and 2 (11%) were CVS+ on EPI but CVS-on SWI (FN), each reflecting absence of a visible vein on SWI. SWI demonstrated high sensitivity of 85% (95% CI: 0.54-0.98) and perfect specificity of 100% (95% CI: 0.48-1), with high accuracy of 89% (95% CI: 0.65-0.99). Agreement for CVS classification was high using AC1 (AC1=0.80, 95% CI: 0.50-1.00, p<0.001), and McNemar’s test was nonsignificant (p=0.5).

All results are provided in Supplementary Table 3.

##### Susceptibility contrast only (EPI magnitude or SWI magnitude)

In Experiment 2 (Figure 1), 5 of 40 lesions (13%) were eligible on both contrasts (TP), 5 lesions (13%) were non-eligible on EPI but eligible on SWI (FP), 20 lesions (50%) were non-eligible on both (TN), and 10 lesions (25%) were eligible on EPI but non-eligible on SWI (FN). All FN lesions were invisible on SWI, and FP lesions again exhibited intralesional veins on EPI that were not visible on SWI.^11^ Notably, raters unanimously judged 29 of the 40 lesions to be non-discernible on SWI, resulting in classification as non-eligible.

SWI demonstrated modest sensitivity of 33% (95% CI: 0.12-0.62) and high specificity of 80% (95% CI: 0.59-0.93). Accuracy was moderate (63%, 95% CI: 0.46-0.77). Agreement was modest but significant (AC1=0.34, 95% CI: 0.02-0.67, p=0.04), with non-significant McNemar’s test (p=0.30).

Among the 5 eligible lesions, 4 (80%) were CVS+ and 1 (20%) was CVS-on both contrasts. No further statistical testing was performed due to insufficient sample size.

All results are provided in Supplementary Table 3.

##### Combination image (FLAIR* or FLAIRswi)

In Experiment 3 (Figure 1), 27 of 40 lesions (68%) were eligible on both contrasts (TP), 1 lesion (3%) was non-eligible on FLAIR* but eligible on FLAIRswi (FP), 11 lesions (28%) were non-eligible on both (TN), and 1 lesion (3%) was eligible on FLAIR* but not FLAIRswi (FN). The FN lesion lacked a visible central vein on FLAIRswi, while the FP lesion showed multiple veins on FLAIR* only.

FLAIRswi demonstrated excellent sensitivity of 96% (95% CI: 0.82-1.00) and excellent specificity of 92% (95% CI: 0.62-1.00), with overall very high accuracy of 95% (95% CI: 0.83-0.99). Eligibility agreement between contrasts was almost perfect, with AC1=0.91 (95% CI: 0.79-1.00, p<0.001). McNemar’s test was nonsignificant (p=1.00).

Among the 27 eligible lesions, 10 (37%) were CVS+ on both contrasts (TP), 10 (37%) were CVS- (TN), and 7 (26%) were CVS+ on FLAIR* but CVS-on FLAIRswi (FN), all lacking a visible central vein on FLAIRswi. FLAIRswi demonstrated moderate sensitivity of 59% (95% CI: 0.32-0.82) and perfect specificity of 100% (95% CI: 0.69-1.00), with accuracy of 74% (95% CI: 0.54-0.89). Agreement for CVS classification was moderate (AC1=0.48, 95% CI: 0.13-0.84, p=0.009). McNemar’s test was significant (p=0.016), reflecting a systematic tendency for SWI to under detect CVS+ lesions compared with EPI.

All results are provided in Supplementary Table 3.

#### PRL Analysis

After re-evaluation of discrepant cases, 27 lesions (23%) out of 120 lesions were rated PRL on both contrasts (TP), 70 lesions (59%) were rated non-PRL on both (TN), 3 lesions (3%) were rated non-PRL on EPI but PRL on SWI (FP), and 20 lesions (17%) were rated PRL on EPI but non-PRL on SWI (FN). Among the 20 FN lesions, 14 (70%) appeared very faint/invisible on SWI, and 6 (30%) showed insufficient differentiation between the hypointense rim and parenchyma. The three FP PRL calls on SWI were attributable to rim-mimicking veins that appeared rim-like on SWI. On EPI, the same findings were recognizable as vascular linear structures and not a true perilesional rim. SWI demonstrated moderate sensitivity of 57% (95% CI: 0.42-0.72) and very high specificity of 95% (95% CI: 0.88-0.99). Overall accuracy was high at 81% (95% CI: 0.73-0.87) and agreement between contrasts was substantial by Gwet’s AC1 (AC1=0.66, 95% CI: 0.52-0.80, p<0.001). McNemar’s test indicated significant directional asymmetry (p<0.001), reflecting a systematic tendency for SWI to under detect PRL relative to EPI.

All results are provided in Supplementary Table 1.

## Discussion

Recent revisions of the McDonald criteria emphasize the diagnostic importance of the CVS and PRL biomarkers.^1^ Although the MAGNIMS-CMSC-NAIMS guidelines provide protocol recommendations,^32^ there is no consensus on which susceptibility-based sequence best supports lesion-level assessment. In this study, SWI demonstrated high specificity for the detection of the CVS and PRL biomarkers, but variable sensitivity and often poor agreement with EPI, influenced by lesion visibility, anatomical location, and the availability of FLAIR information.

We explored differences between EPI and SWI using a whole-brain analysis, which reflects a “real-world” approach simulating what clinicians do in daily practice and a controlled lesion-level analysis, specifically designed to compare individual contrasts or contrast combinations with one another.

The CVS whole brain analysis showed a clear difference in detection performance between the two techniques, with SWI often performing worse in detection of CVS positive lesions than EPI. This imbalance was mostly due to the poor lesion visibility on SWI and, less frequently, to an insufficient contrast between the lesion and its central vein. Additionally, some lesions showed multiple intraparenchymal veins on the EPI which were invisible on the SWI. As highlighted in the recently published guidelines,^32^ this feature (i.e. presence of multiple intralesional veins) represents a lesion exclusion criterion during CVS assessment. Rater feedback provided important context for these findings. In multiple instances, raters reported that intralesional veins could be confirmed on EPI through 3D visualisation when axial views alone were insufficient, whereas comparable vein tracking was not possible on non-isotropic SWI. As stated before, many eligibility discrepancies were attributed to poor lesion parenchymal visibility on SWI magnitude images, which made CVS assessment inherently difficult. Interestingly, some lesions were non-eligible because the lesion parenchyma appeared diffusely hypointense on SWI magnitude images, reducing parenchyma contrast and precluding reliable CVS assessment. After further evaluation, these lesions were identified as PRL on phase imaging.^42,43^ These limitations appeared substantially reduced when using an optimised low-flip angle SWI,^32,46^ with almost no differences in CVS detection performance between the EPI and optimised SWI.

Across the controlled lesion analysis experiments, the performance of SWI consistently improved when the FLAIR anatomical information was available compared to the susceptibility-only imaging scenario, yielding higher sensitivity and stronger κ/AC1 agreement with EPI. In contrast, susceptibility-only evaluations showed markedly lower accuracy and poor reproducibility, attributable to limited parenchymal context and weaker vein - lesion contrast, as noted by the raters. Although eligibility agreement was high in the combination image experiment, CVS classification within eligible lesions remained weaker when using FLAIRswi vs FLAIR*, with more frequent FN ratings on FLAIRswi.

Taken together, these results highlight that the FLAIR-based anatomical context is critical for a reliable CVS assessment, with SWI contrast alone being often insufficient for a robust CVS biomarker evaluation. Overall, our findings align with prior work demonstrating superior venous conspicuity on T2*-weighted EPI across field strengths,^22-24^ as well as meta-analytic evidence showing higher CVS positive lesions proportions on high-resolution EPI relative to SWI.^20,25^ In line with the recent NAIMS guidelines,^32^ our single-case example suggests that optimized SWI, particularly with lower flip angles,^46^ improves both venous and lesion visibility in MS. This supports current recommendations suggesting to refine SWI acquisition parameters rather than relying on conventional SWI implementations.^32,46^

Regarding the PRL assessment, the discrepancies in biomarker detection performance appeared overall attenuated, with substantial/good agreement between EPI and SWI (using phase- and magnitude-based susceptibility images with anatomical reference). However, McNemar’s test demonstrated significant directional disagreement for both the whole brain and the controlled lesion analysis, indicating a systematic tendency of the SWI contrast to under detect PRL relative to EPI. False negatives typically involved lesions that were partially invisible on SWI, failed to meet the two-thirds rim requirement, or were located in regions with strong SWI susceptibility gradients such as the optic pathways and occipital horn. False positives were rare and largely attributable to veins mimicking rims or, especially for infratentorial lesions or lesions located in the temporal lobe due to susceptibility-induced geometric distortions and signal dropout on EPI.^44,45^ Importantly, these findings also indicate that rim visibility is region-dependent for both contrasts, with SWI and EPI each showing vulnerability to susceptibility-related artifacts in specific anatomical regions.

However, when considered across lesion locations and routine clinical image quality, SWI appears to be more affected by heterogeneity in image quality, local susceptibility,^47^ and rim morphology under routine clinical conditions. Some discrepancies may also reflect pseudo-rims or dipolar phase artifacts previously described in susceptibility-based imaging. ^48,49^

### Limitations

In all whole-brain analyses, lesions that were negative on both contrasts were excluded. This removed true negatives by design and introduced substantial prevalence imbalance, which inflates sensitivity and precision while markedly suppressing κ, a statistic highly dependent on class proportions, whereas AC1 provides a complementary estimate that is less sensitive to prevalence imbalance but should be interpreted alongside κ and McNemar’s results rather than as a replacement. Agreement metrics should therefore be interpreted with these constraints in mind. Throughout this study, high-resolution 3D T2*-EPI was used as an operational comparator rather than a true gold standard, acknowledging that both EPI and SWI are affected by sequence-specific susceptibility artifacts that may differentially impact lesion-level classification. Indeed, limitations include sequence-specific artifacts, rater variability, and the possibility that lesion distributions were not perfectly matched across controlled lesion analysis sub-experiments represent additional limitations. Finally, imaging differences cannot be fully separated from underlying biological variation in venous visibility or rim morphology, which may also contribute to classification differences between SWI and EPI.

## Conclusion

Across both CVS and PRL analyses, SWI diverged systematically from EPI, particularly in whole-brain settings where reduced lesion and vein visibility on SWI limited lesion-level reliability. While EPI cannot be considered an absolute gold standard, the consistent direction and magnitude of disagreement indicate that standard clinical SWI should be used with caution for CVS or PRL classification.

As CVS and PRL assessment increasingly informs differential diagnosis and prognostic stratification in MS, inaccuracies introduced by conventional SWI may affect clinical decision-making, especially in patients with low lesion burden or atypical presentations. Importantly, in line with recent studies,^32,46^ our findings also suggest that optimised SWI acquisition, such as those employing lower flip angles, can substantially improve lesion and venous conspicuity. These observations support ongoing effort to refine SWI acquisition parameters, rather than relying on standard implementations, to enable more reliable CVS and PRL assessment in clinical practice.

## Data Availability

All data produced in the present study are available upon reasonable request to the authors. Data will be shared upon establishment of a data-sharing agreement with formal approval from the local ethics committee.

## Acknowledgements

The authors thank the study participants; the neuroimmunology clinics of cliniques universitaires Saint-Luc (CUSL, Brussels, Belgium) and NINDS (NIH Clinical Center, Bethesda, Maryland, USA) for recruiting and evaluating the patients and for coordinating the scans; Thierry Duprez, Sébastien de Laever (CUSL), Laurence Dricot (Université catholique de Louvain, Oceane Perdaens and Julie Poujol (GE Healthcare) for assistance with 3T MRI scan acquisition and analysis in Brussels.

## Disclosures

Anna Stölting: has the financial support of the Fédération Wallonie Bruxelles – FRIA du Fonds de la Recherche Scientifique – FNRS.

Eline van Doninck: is supported by FWO-TBM (project MACSiMiSE-BRAIN—T001121N)

Serena Borrelli: is supported by the Funds Claire Fauconnier, Ginette Kryksztein & José and Marie Philippart-Hoffelt, managed by the King Baudouin Foundation. She has received unrelated research funding from Roche, Sanofi, and Brugmann Foundation.

Colin Vanden Bulcke: nothing to disclose.

Maria Sofia Martire: nothing to disclose.

Francois Guisset: nothing to disclose.

Maxence Wynen: nothing to disclose.

Gaetan Duchene: was employed by GE Healthcare untill January 2023 as an MR scientist on belgian academic sites, including Cliniques Universitaires Saint-Luc.

Lucia Moiola: received honoraria for speaking and for partecipating to advisory board from Merck, Celgene, Biogen, Sanofi, Novartis, Roche, Alexion, Amgen, Neuraxpharm

Veronica Popescu: has received honoraria and travel and research grants from the Research Foundation - Flanders (FWO), Almirall, Biogen, BMS, Janssen Pharmaceutica, Merck, Neuraxpharma, Roche, Sanofi-Genzyme, Viatris.

Barbara Willekens: received travel support to attend conferences and meetings from Biogen, Sanofi-Genzyme, Merck, Neuraxpharm, Novartis, Roche, UCB, EAN, European Charcot Foundation, ECTRIMS, CMSC, MEDEN. BW has acted as a consultant during scientific advisory boards and/or as speaker for Alexion, Almirall, Biogen, BMS-Celgene, Galapagos, Horizon, Immunic Therapeutics, Janssen, Merck, Novartis, Roche, Sandoz, Sanofi-Genzyme. All fees were paid to UZA. BW has received research and/or patient support grants and/or sponsoring for educational events from UZA Foundation, Belgian Charcot Foundation, Start2Cure Foundation, Research Foundation (FWO) Flanders, OJO, Horlait-Dapsens Medical Foundation, EJP-RD Networking Support Scheme, Queen Elisabeth Medical Foundation, National MS Society USA, Amgen, Biogen, Janssen, Sanofi-Genzyme, Novartis, Merck, Roche. All funds/grants were paid to UZA Foundation, UZA or UA.

Massimo Filippi: is Editor-in-Chief of the Journal of Neurology, Associate Editor of Human Brain Mapping, Neurological Sciences, and Radiology; received compensation for consulting services from Alexion, Almirall, Biogen, Merck, Novartis, Roche, Sanofi; speaking activities from Bayer, Biogen, Celgene, Chiesi Italia SpA, Eli Lilly, Genzyme, Janssen, Merck-Serono, Neopharmed Gentili, Novartis, Novo Nordisk, Roche, Sanofi, Takeda, and TEVA; participation in Advisory Boards for Alexion, Biogen, Bristol-Myers Squibb, Merck, Novartis, Roche, Sanofi, Sanofi-Aventis, Sanofi-Genzyme, Takeda; scientific direction of educational events for Biogen, Merck, Roche, Celgene, Bristol-Myers Squibb, Lilly, Novartis, Sanofi-Genzyme; he receives research support from Biogen Idec, Merck-Serono, Novartis, Roche, the Italian Ministry of Health, the Italian Ministry of University and Research, and Fondazione Italiana Sclerosi Multipla. M.A. received speaker and/or consulting honoraria from Biogen, Sanofi, Abata Therapeutics, GSK and Immunic Therapeutics.

Martina Absinta: received research support from the European Research Council, National MS Society, Conrad N Hilton Foundation, International Progressive MS Alliance, Cariplo Foundation, FRRB Early Career Award, and FISM - Fondazione Italiana Sclerosi Multipla co-financed with the ‘5 per mille’ public funding. MA also received consultancy and/or speaker honoraria from Sanofi, GSK, Biogen, Roche and Immunic Therapeutics.

Pietro Maggi: is supported by the Fondation Charcot Stichting Research Fund 2023 and 2025, the Fund for Scientific Research (F.R.S, FNRS; grant #40008331), Cliniques universitaires Saint-Luc “Fonds de Recherche Clinique”, Biogen and Sanofi. He has received consulting honoraria from Sanofi, Biogen, Merck and Novartis.

## Notes

### Competing Interest Statement

The authors have declared no competing interest.

### Funding Statement

AS has financial support from the Federation Wallonie Bruxelles and FRIA du Fonds de la Recherche Scientifique (FNRS).
EvDo is supported by FWO-TBM (project MACSiMiSE-BRAIN-T001121N).
SB is supported by the Funds Claire Fauconnier, Ginette Kryksztein, and Jose and Marie Philippart-Hoffelt, managed by the King Baudouin Foundation. She has received unrelated research funding from Roche, Sanofi, and the Brugmann Foundation.
CVB has nothing to disclose.
MSM has nothing to disclose.
FG has nothing to disclose.
MW has nothing to disclose.
GD was employed by GE Healthcare until January 2023 as an MR scientist on Belgian academic sites, including Cliniques Universitaires Saint-Luc.
LM received honoraria for speaking and for participating in advisory boards from Merck, Celgene, Biogen, Sanofi, Novartis, Roche, Alexion, Amgen, and Neuraxpharm.
VP has received honoraria and travel and research grants from the Research Foundation Flanders (FWO), Almirall, Biogen, BMS, Janssen Pharmaceutica, Merck, Neuraxpharma, Roche, Sanofi-Genzyme, and Viatris.
BW received travel support to attend conferences and meetings from Biogen, Sanofi-Genzyme, Merck, Neuraxpharm, Novartis, Roche, UCB, EAN, the European Charcot Foundation, ECTRIMS, CMSC, and MEDEN. BW has acted as a consultant during scientific advisory boards and or as a speaker for Alexion, Almirall, Biogen, BMS-Celgene, Galapagos, Horizon, Immunic Therapeutics, Janssen, Merck, Novartis, Roche, Sandoz, and Sanofi-Genzyme. All fees were paid to UZA. BW has received research and or patient support grants and or sponsorship for educational events from the UZA Foundation, Belgian Charcot Foundation, Start2Cure Foundation, Research Foundation Flanders (FWO), OJO, Horlait-Dapsens Medical Foundation, EJP-RD Networking Support Scheme, Queen Elisabeth Medical Foundation, National MS Society USA, Amgen, Biogen, Janssen, Sanofi-Genzyme, Novartis, Merck, and Roche. All funds and grants were paid to the UZA Foundation, UZA, or UA.
MF is Editor-in-Chief of the Journal of Neurology and Associate Editor of Human Brain Mapping, Neurological Sciences, and Radiology. He received compensation for consulting services from Alexion, Almirall, Biogen, Merck, Novartis, Roche, and Sanofi, and for speaking activities from Bayer, Biogen, Celgene, Chiesi Italia SpA, Eli Lilly, Genzyme, Janssen, Merck-Serono, Neopharmed Gentili, Novartis, Novo Nordisk, Roche, Sanofi, Takeda, and TEVA. He participated in advisory boards for Alexion, Biogen, Bristol-Myers Squibb, Merck, Novartis, Roche, Sanofi, Sanofi-Aventis, Sanofi-Genzyme, and Takeda. He was involved in the scientific direction of educational events for Biogen, Merck, Roche, Celgene, Bristol-Myers Squibb, Lilly, Novartis, and Sanofi-Genzyme. He receives research support from Biogen Idec, Merck-Serono, Novartis, Roche, the Italian Ministry of Health, the Italian Ministry of University and Research, and Fondazione Italiana Sclerosi Multipla.
MA received speaker and or consulting honoraria from Biogen, Sanofi, Abata Therapeutics, GSK, and Immunic Therapeutics. She also received research support from the European Research Council, National MS Society, Conrad N Hilton Foundation, International Progressive MS Alliance, Cariplo Foundation, FRRB Early Career Award, and FISM Fondazione Italiana Sclerosi Multipla, co-financed with the 5 per mille public funding. MA also received consultancy and or speaker honoraria from Sanofi, GSK, Biogen, Roche, and Immunic Therapeutics.
PM is supported by the Fondation Charcot Stichting Research Fund 2023 and 2025, the Fund for Scientific Research (FRS FNRS; grant 40008331), the Cliniques universitaires Saint-Luc Fonds de Recherche Clinique, Biogen, and Sanofi. He has received consulting honoraria from Sanofi, Biogen, Merck, and Novartis.

### Author Declarations

Clinical and imaging data were collected under institutional review board-approved protocols in two university hospitals (Cliniques universitaires Saint-Luc in Brussels, Belgium and Multiple Sclerosis Center, IRCCS San Raffaele Hospital in Milan, Italy).

