## Supplementary Material for "Comparative evaluation of EPI and SWI for the assessment of PRL and CVS in Multiple Sclerosis"

### Supplementary Materials

**Image Analysis**

Analyses were conducted in two independent components: whole-brain assessments and controlled lesion-based assessments. An overview of the experimental workflow can be found in Figure 1.

***Whole-Brain Assessment***

Whole-brain images were evaluated by experienced raters to assess the PRL status and CVS of all FLAIR visible lesions on both EPI and SWI images. Anonymized images from both centres were randomly mixed.

*CVS*

Two experienced raters (F.G. and P.M.) independently rated the CVS on EPI and SWI, respectively Experiment 1 and 2, here below. Experiment 2 was performed ≥4 weeks after Experiment 1 to ensure blinding.

- **Experiment 1:** EPI magnitude + registered FLAIR
- **Experiment 2:** SWI magnitude + registered FLAIR (≥4 weeks later)

Raters annotated CVS+ and CVS- lesions using ITK-Snap. Disagreements were adjudicated by two independent experts (S.B. and A.S.), and final consensus masks were created.

*PRL*

As for the CVS, two experienced raters (A.S. and E.v.D.) evaluated PRL status on both EPI and SWI (respectively Experiment 1 and 2, here below), and Experiment 2 was always performed ≥4 weeks apart.

- **Experiment 1:** filtered EPI phase + EPI magnitude + FLAIR
- **Experiment 2:** SWI phase + SWI magnitude + FLAIR (≥4 weeks later)

Disagreements were resolved by a third rater (P.M.), producing final consensus masks.

***Controlled Lesion Analysis Assessment***

A set of 120 lesions was pre-selected and extracted into standardized PDFs. Lesion selection was performed separately for PRL and CVS assessments, based on modality-specific visibility criteria (PRL: A.S. and E.v.D.; CVS: A.S.). For each assessment, disagreements were resolved by expert adjudication (PRL: P.M. and A.S.; CVS: S.B. and A.S.). Lesions were cropped around manually identified centres (40×40 voxels), and brightness/contrast were optimized per subject and contrast.

*CVS*

Three experienced raters (M.A., S.B. and M.S.M.) independently evaluated the CVS in 120 lesions using EPI-based susceptibility imaging. Lesions were equally distributed across three imaging contrast scenarios (40 lesions per scenario):

- **Experiment 1A:** FLAIR + susceptibility contrast
- **Experiment 2A:** Susceptibility contrast only
- **Experiment 3A:** FLAIR* or FLAIRswi

After ≥4 weeks, CVS assessment for the same lesions was repeated using the SWI contrast (instead of the EPI) while preserving the same three imaging contrast scenarios (Experiments 1B, 2B, and 3B).

*PRL*

A separate set of 120 lesions was evaluated by trained raters (M.A., P.M., M.S.M., V.P., M.W. and B.W.).

- Round 1: filtered EPI phase + EPI magnitude + FLAIR + MPRAGE
- Round 2: SWI phase + SWI magnitude + FLAIR + MPRAGE (≥4 weeks later)

PRL and CVS Assessment Criteria

PRLs were assessed according to the North American Imaging in Multiple Sclerosis (NAIMS) guidelines^1^ based on following criteria:

- A discrete, hypointense, continuous paramagnetic rim covering at least two-thirds of the lesion’s outer edge
- Rim colocalization with the edge of a hyperintense core on T2-weighted images, or in the case of large lesions, with a hypointense core on T1-weighted images
- Visibility of the rim on at least two consecutive slices (for 2D acquisitions) or in two orthogonal planes (for 3D acquisitions)
- Absence of colocalization with gadolinium-enhancing lesions

The central vein sign (CVS) was also evaluated per NAIMS criteria^2^ as follows:

*Inclusion criteria*

- A thin hypointense line or a small hypointense dot within the lesion
- Visibility in at least two perpendicular MRI planes (3D) or two consecutive slices (2D)
- Central location within the lesion
- Vein diameter < 2mm

*Exclusion criteria*

- Lesions smaller 3mm in any plane
- Confluent lesions
- Lesions with multiple veins

MRI Protocols

Participants at Saint Luc University Hospital underwent brain 3T-MRI (GE SIGNA^TM^ Premier research scanner, General Electric, Milwaukee, WI) equipped with a 48-channel head coil. The MRI protocol included
(**i**) sagittal 3D T1-weighted magnetization-prepared rapid gradient echo (MPRAGE) sequence (repetition time (TR)=2186 ms, echo-time (TE)=3 ms, inversion time (TI)=900 ms, number of slices (#slices)=156, voxel size=1.0x1.0x1.0 mm^3^),
(**ii**) sagittal 3D FLAIR sequence (TR=5000ms, TE=105ms, TI=1532 ms, #slices=340, voxel size=1.0x1.0x1.0 mm^3^),
(**iii**) a sagittal high-resolution 3D T2*-weighted echo-planar imaging (EPI)^3^ sequence (TR=80.2 ms, TE=35 ms, flip angle = 18°, #slices=355, voxel size=0.67x0.67x0.67 mm^3^),
(**iv**) a 3D axial susceptibility sequence (GE SWAN) providing T2*-magnitude and unwrapped filtered phase contrast was acquired (TR = 50 ms, TE = 20.0 ms, flip angle = 15°, #slices = 96, acquisition voxel size = 0.5x0.5x1.8 mm^3^)
(**v**) a low-flip-angle adapted (GE SWAN) providing T2*-magnitude and unwrapped filtered phase contrast was acquired (TR = 36.5 ms, TE = 25.6 ms, flip angle = 12°, #slices = 240, acquisition voxel size = 0.75x0.75x1.0 mm^3^)
(**vi**) a post-gadolinium T1-weighted spoiled gradient recalled echo (CE-SPGR) sequence (TR=6.9 ms, TE=2.1 ms, flip angle=12°, #slices=150, voxel size=0.75x0.75x1.0 mm^3^).

Participants in San Raffaele Hospital, Milan underwent brain 3T MRI (Philips Ingenia 3T CX) equipped with a 32-channel head coil. The MRI protocol included:
(**i**) a axial 3D T1-weighted magnetization-prepared rapid gradient echo sequence (Philips T1-TFE) ; TR = 9.4 ms, TE = 4.6 ms, flip angle = 8°, #slices = 50, voxel size = 0.24x0.24x3 mm^3^),
(**ii**) a whole-brain axial 3D T2-weighted FLAIR sequence (TR = 4800 ms, TE = 373 ms, TI = 1600ms, flip angle = 90°, #slices = 70, voxel size = 0.47 × 0.47 × 2 mm^3^),
(**iii**) an axial high-resolution 3D T2*-weighted echo-planar imaging (EPI) sequence (TR = 50.7ms, TE = 27.7ms, flip angle = 10°, #slices = 336, voxel size = 0.5x0.5x0.55 mm^3^), and
(**iv**) a whole-brain 3D axial susceptibility-weighted imaging (SWI) sequence providing T2*-magnitude and unwrapped filtered phase contrasts (TR = 31 ms, TE = 7.2 ms, flip angle = 17°, #slices = 261, acquisition voxel size = 0.45 × 0.45 × 1 mm^3^).

**Supplementary Tables**

Table 1 - Agreement metrics for PRL controlled lesion and whole brain analyses

|  | PRL Whole Brain Analysis | | PRL Controlled Lesion Analysis | |
| --- | --- | --- | --- | --- |
| **Metric** | **Results** | **p-value** | **Results** | **p-value** |
| Total lesions observed | 311 |  | 120 |  |
| True Positives (TP) | 265 (85.2%) |  | 27 |  |
| True Negatives (TN) | 0 (0%) |  | 70 |  |
| False Positives (FP) | 9 (2.9%) |  | 3 |  |
| False Negatives (FN) | 37 (11.9%) |  | 20 |  |
| Sensitivity | 0.877 [0.835, 0.912] | p<0.0001 | 0.574 [0.422, 0.717] | 0.382 |
| Specificity | 0.000 [0.000, 0.336] | 0.004 | 0.959 [0.885, 0.991] | p<0.0001 |
| Accuracy | 0.852 [0.808, 0.890] | p<0.0001 | 0.808 [0.726, 0.874] | p<0.0001 |
| Precision | 0.967 [0.939, 0.985] | p<0.0001 | 0.900 [0.735, 0.979] | p<0.0001 |
| PPV | 0.967 [0.939, 0.985] | p<0.0001 | 0.900 [0.735, 0.979] | p<0.0001 |
| NPV | 0.000 [0.000, 0.095] | p<0.0001 | 0.778 [0.678, 0.859] | p<0.0001 |
| LR+ | 0.922 [0.795, 1.070] | 0.286 | 13.979 [4.493, 43.494] | p<0.0001 |
| LR− | 2.475 [0.163, 37.507] | 0.513 | 0.444 [0.317, 0.621] | p<0.0001 |
| F1 Score | 0.920 |  | 0.701 |  |
| Cohen’s κ | -0.049 [-0.329, 0.231] | 0.263 | 0.570 [0.412, 0.728] | p<0.0001 |
| Gwet’s AC1 | 0.829 [0.776, 0.881] | p<0.0001 | 0.660 [0.523, 0.797] | p<0.0001 |
| McNemar Test | χ² = 15.848 | p<0.0001 | χ² = 11.130 | p<0.0001 |

Table 2 -Agreement Metrics for CVS whole brain analyses with eligibility (CVS eligible vs non-eligible) and lesion positivity (CVS+ vs CVS-)

|  | CVS Whole Brain Analysis Eligibility | | CVS Whole Brain Analysis Lesion Positivity | |
| --- | --- | --- | --- | --- |
| **Metric** | **Results** | **p-value** | **Results** | **p-value** |
| Total lesions observed | 174 |  | 131 |  |
| True Positives (TP) | 131 (75.4%) |  | 104 |  |
| True Negatives (TN) | 0 (0%) |  | 4 |  |
| False Positives (FP) | 6 (3.4%) |  | 0 |  |
| False Negatives (FN) | 37 (21.1%) |  | 23 |  |
| Sensitivity | 0.78 [0.709, 0.84] | p<0.0001 | 0.819 [0.741, 0.882] | p<0.0001 |
| Specificity | 0.000 [0.000, 0.459] | 0.031 | 1 [0.398, 1] | 0.125 |
| Accuracy | 0.752 [0.682, 0.815] | p<0.0001 | 0.824 [0.748, 0.885] | p<0.0001 |
| Precision | 0.956 [0.907, 0.984] | p<0.0001 | 0.981 [0.934, 0.998] | p<0.0001 |
| PPV | 0.956 [0.907, 0.984] | p<0.0001 | 1 [0.965, 1] | p<0.0001 |
| NPV | 0.000 [0.000, 0.095] | p<0.0001 | 0.148 [0.042, 0.337] | 0.0003 |
| LR+ | 0.838 [0.673, 1.046] | 0.116 | 8.164 [0.588, 113.363] | 0.118 |
| LR− | 3.107 [0.212, 45.575] | 0.408 | 0.203 [0.128, 0.326] | p<0.0001 |
| F1 Score | 0.859 |  | 0.900 |  |
| Cohen’s κ | -0.063 [-0.108, -0.018] | 0.195 | 0.216 0.036, 0.397] | p<0.0001 |
| Gwet’s AC1 | 0.685 [0.582, 0.787] | p<0.0001 | 0.778 [0.681, 0.875] | p<0.0001 |
| McNemar Test | χ² = 20.930 | p<0.0001 | χ² = 21.043 | p<0.0001 |

Table 3 - Agreement Metrics for CVS whole brain analyses with eligibility (CVS eligible vs non-eligible) and lesion positivity (CVS+ vs CVS-). Experiment 1 includes FLAIR + susceptibility contrast, Experiment 2 susceptibility contrast only and Experiment 3 the fused FLAIR*/FLAIRswi

| Metric | Experiment 1 Eligibility | Experiment 1 Eligibility p value | Experiment 1 Positivity | Experiment 1 Positivity p value | Experiment 2 Eligibility | Experiment 2 Eligibility p value | Experiment 2 Positivity | Experiment 2 Positivity p value | Experiment 3 Eligibility | Experiment 3 Eligibility p value | Experiment 3 Positivity | Experiment 3 Positivity p value |
| --- | --- | --- | --- | --- | --- | --- | --- | --- | --- | --- | --- | --- |
| Total lesions (Eligibility) | 40 |  | 18 |  | 40 |  | 5 |  | 40 |  | 27 |  |
| True Positives (TP) | 18 (45%) |  | 11 |  | 5 (12.5%) |  | 4 |  | 27 (67.5%) |  | 10 |  |
| True Negatives (TN) | 14 (35%) |  | 5 |  | 20 (50%) |  | 1 |  | 11 (27.5%) |  | 10 |  |
| False Positives (FP) | 3 |  | 0 |  | 5 |  | 0 |  | 1 |  | 0 |  |
| False Negatives (FN) | 5 |  | 2 |  | 10 |  | 0 |  | 1 |  | 7 |  |
| Sensitivity | 0.783 [0.563, 0.925] | 0.0106 | 0.846 [0.546, 0.981] | 0.0225 | 0.333 [0.118, 0.616] | 0.3018 | 1.000 [0.398, 1.000] | 0.1250 | 0.964 [0.817, 0.999] | p<0.0001 | 0.588 [0.329, 0.816] | 0.6291 |
| Specificity | 0.824 [0.566, 0.962] | 0.0127 | 1.000 [0.478, 1.000] | 0.0625 | 0.800 [0.593, 0.932] | 0.0041 | 1.000 [0.025, 1.000] | 10.000 | 0.917 [0.615, 0.998] | 0.0063 | 1.000 [0.692, 1.000] | 0.0020 |
| Accuracy | 0.800 [0.644, 0.909] | 0.0002 | 0.889 [0.653, 0.986] | 0.0013 | 0.625 [0.458, 0.773] | 0.1539 | 1.000 [0.478, 1.000] | 0.0625 | 0.950 [0.831, 0.994] | p<0.0001 | 0.741 [0.537, 0.889] | 0.0192 |
| Precision | 0.857 [0.637, 0.970] | 0.0015 | 1.000 [0.715, 1.000] | 0.0010 | 0.500 [0.187, 0.813] | 1 | 1.000 [0.398, 1.000] | 0.1250 | 0.964 [0.817, 0.999] | p<0.0001 | 1.000 [0.692, 1.000] | 0.0020 |
| PPV | 0.857 [0.637, 0.970] | 0.0015 | 1.000 [0.715, 1.000] | 0.0010 | 0.500 [0.187, 0.813] | 1 | 1.000 [0.398, 1.000] | 0.1250 | 0.964 [0.817, 0.999] | p<0.0001 | 1.000 [0.692, 1.000] | 0.0020 |
| NPV | 0.737 [0.488, 0.909] | 0.0636 | 0.714 [0.290, 0.963] | 0.4531 | 0.667 [0.472, 0.827] | 0.0987 | 1.000 [0.025, 1.000] | 10.000 | 0.917 [0.615, 0.998] | 0.0063 | 0.588 [0.329, 0.816] | 0.6291 |
| LR+ | 4.435 [1.553, 12.663] | 0.0054 | 9.857 [0.686, 141.630] | 0.0924 | 1.667 [0.577, 4.818] | 0.3456 | 3.600 [0.321, 40.411] | 0.2992 | 11.571 [1.769, 75.673] | 0.0106 | 12.833 [0.832, 197.972] | 0.0675 |
| LR− | 0.264 [0.118, 0.591] | 0.0012 | 0.195 [0.062, 0.615] | 0.0053 | 0.833 [0.554, 1.253] | 0.3811 | 0.133 [0.009, 2.083] | 0.1508 | 0.039 [0.006, 0.269] | 0.0010 | 0.437 [0.249, 0.765] | 0.0038 |
| F1 Score | 0.818 |  | 0.917 |  | 0.400 |  | 1.000 |  | 0.964 |  | 0.741 |  |
| Cohen’s κ | 0.597 [0.347, 0.847] | 0.0001 | 0.753 [0.431, 1.076] | 0.0010 | 0.143 [-0.200, 0.486] | 0.3458 | 1.000 [1.000, 1.000] | 0.0253 | 0.881 [0.720, 1.042] | p<0.0001 | 0.514 [0.204, 0.824] | 0.0022 |
| Gwet’s AC1 | 0.604 [0.345, 0.863] | p<0.0001 | 0.800 [0.498, 1.000] | p<0.0001 | 0.342 [0.015, 0.670] | 0.0410 | 1.000 [1.000, 1.000] | p<0.0001 | 0.914 [0.789, 1.000] | p<0.0001 | 0.481 [0.128, 0.835] | 0.0095 |
| McNemar Test | χ² = 0.125 | 0.7297 | χ² = 0.500 | 0.4795 | χ² = 1.067 | 0.3017 | χ² = NaN |  | χ² = 0.000 | 1 | χ² = 5.143 | 0.0156 |
